# Reliability and Stability of Cerebral Palsy Classification Scales for Individuals with *STXBP1* Related Disorders and *SYNGAP1* Related Disorders

**DOI:** 10.1101/2025.11.04.25339413

**Authors:** Samuel R. Pierce, Julie M. Orlando, Kristin G. Cunningham, Sarah M. Ruggiero, Jillian L. McKee, Ingo Helbig

## Abstract

**Aim:** To determine the interrater reliability and stability of the Gross Motor Function Classification System (GMFCS), mini–Manual Ability Classification System (mini-MACS), Manual Ability Classification System (MACS), and Communication Function Classification System (CFCS) in individuals with *STXBP1*-Related Disorder (*STXBP1*-RD) and *SYNGAP1*-Related Disorder (*SYNGAP1*-RD).

**Methods:** Data were collected from 83 individuals with *STXBP1*-RD (mean age = 9.8 years) and 101 individuals with *SYNGAP1*-RD (mean age = 10.9 years). Two raters completed the GMFCS, MACS/MiniMACS, and CFCS assessments on the same day, and test-retest stability was evaluated for participants with two longitudinal assessments.

**Results:** Interrater agreement varied from 73.8% to 77.3% for the *STXBP1*-RD cohort and from 60.5% to 83.3% for the *SYNGAP1*-RD cohort. Interrater reliability weighted kappa scores for the *STXBP1*-RD cohort varied from 0.83 to 0.93 while the *SYNGAP1*-RD cohort ranged from 0.66-0.81. Test-retest stability scores for the *STXBP1*-RD group varied from 0.62 to 0.94 while the *SYNGAP1*-RD group ranged from 0.38 to 0.78. Significant correlations were found between all assessment scales for both *STXBP1*-RD (Kendall’s Tau range from 0.25-0.42) and *SYNGAP1*-RD (Kendall’s Tau range from 0.19-0.45).

**Interpretation:** The GMFCS, MACS/MiniMACS, and CFCS demonstrate appropriate levels of interrater reliability and stability for individuals with *STXBP1*-RD and *SYNGAP1*-RD.

**What this paper adds:**

- Classification tools are reliable and stable in individuals with *STXBP1*-RD and *SYNGAP1*-RD.
- Gross motor function is least affected for both conditions.
- Language function is most affected for both conditions.
- Correlations are decreased compared to children with cerebral palsy due to phenotype differences.

## Background

Access to genetic testing and counseling has increased dramatically in the past decade, as genetic diagnostic technologies are increasingly seen as standard-of-care for individuals with neurodevelopmental disorders and costs of technology continue to decrease.^1.2^ Exome and genome sequencing for children with developmental delays and intellectual disabilities are recommended by the American College of Medical Genetics and Genomics as first line tests to potentially determine the cause of the child’s presentation.^3^ As a result, the diagnosis of rare neurological conditions has become more common, which may improve clinical care and allow for the development of more precise treatments.^4^ Accordingly, it is reasonable to expect that pediatric healthcare providers can expect that these collectively rare conditions will become a more common area of practice as more individuals are diagnosed. Accordingly, using valid and reliable outcome measures in this patient population for routine clinical and emerging precision medicine trials is increasingly important.^5^

*STXBP1*-Related Disorders (*STXBP1*-RD) and *SYNGAP1*-Related Disorders (*SYNGAP1*-RD) are two individually rare diagnoses, yet are two of the most commonly identified genetic causes of neurodevelopmental disorders and epilepsy.^6^ In 2008, *STXBP1* was first discovered as a disease gene when pathogenic variants were identified in 5 individuals with Ohtahara syndrome, an early-onset epilepsy associated with severe developmental delays.^7^ However, the known phenotypic range of *STXBP1*-RD has been growing. The largest cohort of individuals with *STXBP1*-RD was described in Xian et al.^8^ This study assessed phenotypes in 534 individuals with *STXBP1*-RD and identified global developmental delay, absent speech, and seizures as common features of this condition. Furthermore, qualitative interviews of caregivers and providers of individuals with *STXBP1*-RD performed by Sullivan et al showed that difficulties with gross motor function, expressive communication, and autonomy are common concerns for many families.^9^

The *SYNGAP1* gene was identified in 2009^10^ and first recognized as a cause of developmental and epileptic encephalopathy in 2013.^11^ An investigation by Wright et al^12^ found that parents and caregivers of individuals with *SYNGAP1*-RD reported limitations in fine and gross motor skills as well as high rates of autism spectrum disorder and epilepsy. A recent paper by McKee et al reported that phenotypes associated with *SYNGAP1-RD* include behavioral challenges, generalized-onset seizures, autism spectrum disorder, and prominent language deficits while gross motor and fine motor skill acquisition are often delayed.^13^

While both *STXBP1*-RD and *SYNGAP1*-RD are becoming more commonly diagnosed and are targets for drug and gene therapy,^14,15^ there is limited information describing the gross motor, fine motor, and language function of individuals with these conditions. However, due to the rarity of these disorders, utilizing existing clinical assessments for diagnoses with a similar presentation such as cerebral palsy (CP) may be necessary due to the resources needed to develop new condition-specific outcome measures. Clinical assessments developed for children with CP may also be valid for use with individuals with *STXBP1*-RD and *SYNGAP1*-RD because these conditions fall under the umbrella diagnosis of CP.^16^

One potential set of tools that may be useful in describing function in individuals with *STXBP1*-RD and *SYNGAP1*-RD are the functional classification scales developed for children with CP. The Gross Motor Function Classification System (GMFCS)^17^, mini–Manual Ability Classification System (mini-MACS)^18^, Manual Ability Classification System (MACS)^19^, and Communication Function Classification System (CFCS)^20^ are five-point ordinal scales used to describe gross motor, fine motor, and communication function respectively with lower scores indicating better function. Xian et al reported variable GMFCS and MACS scores in individuals with *STXBP1*-RD while more severe impairments in CFCS scores were evident.^21^ A recent investigation by Thalwitzer et al^22^ found that GFMCS scores were higher in individuals with *STXBP1*-RD with seizures compared to those without seizures. To date, there are no studies using the GMFCS, MACS/miniMACS, or CFCS in individuals with *SYNGAP1*-RD. Significant correlations between these scales have also been reported in children with CP.^23^ However, it remains unknown if similar correlations are present in individuals with *STXBP1*-RD and *SYNGAP1*-RD since their clinical phenotypes may be different. While the reliability^24^ and stability^25^ of these scales has been well studied in the children with CP, the reliability and stability of these classification systems has not been reported to date in rare genetic conditions such as *STXBP1*-RD and *SYNGAP1*-RD. The aims of this investigation are to:

1. Describe the gross motor, fine motor, and language function using the GMFCS, MACS, mini-MACS, or CFCS of a large cohort of individuals with *STXBP1*-RD and *SYNGAP1*-RD and determine the relations between these functional scales.
2. Determine the interrater reliability and stability of the GMFCS, MACS/mini-MACS, and CFCS in individuals with *STXBP1*-RD and *SYNGAP1*-RD

## Methods

Participants were recruited as part of a prospective natural history study of *SYNGAP1*-RD and *STXBP1-*RD was approved by the Institutional Review Board of Children’s Hospital of Philadelphia. Parents or legal guardians of all participants provided written, informed consent. Participants met the following inclusion criteria: a diagnosis of *SYNGAP1*-RD or *STXBP1*-RD as determined by genetic testing as causative based on clinical and variant classification criteria. Exclusion criterion were: 1) a confirmed variant in a gene other than *SYNGAP1*-RD *or STXBP1*-RD that is known to contribute to a neurodevelopmental disability; 2) a significant non-*SYNGAP1*-RD or non-*STXBP1*-RD related central nervous impairment; 3) History of prematurity defined as gestational age <34 weeks, interventricular hemorrhage, structural brain deficit or congenital heart disease. Participants were asked to complete an assessment visit every 6 months, however, some participants completed assessment visits annually due to travel barriers or family preference.

### Procedure

Raters were trained prior to data collection on the classification scales. The GMFCS was completed in all participants. The miniMACS was completed in subjects between the ages of 1 and 4 years while the MACS was used in subjects over the age of 4. The CFCS was completed in all participants at least 2 years of age as per scale guidelines. While the CFCS, GMFCS, and MACS were designed for children under the age of 18, participants over the age of 18 were assessed using these scales as per the natural history study protocol which was designed to assess both adults and children with *STXBP1*-RD or *SYNGAP1*-RD.

### Functional Assessment Scale Assessment

Two assessors completed the GMFCS, MACS/MiniMACS, and CFCS assessments on the same day. Scales were independently scored via direct observation of skills, parent interview, or a combination of both observation and parent interview which occurred during physical therapy and occupational therapy examinations. Disagreements in scoring were discussed between the assessors, and a consensus score was determined which was used as the score for each scale. We then evaluated the relations between scores on the GMFCS, MACS, and CFCS assessments at the first assessment visit for each participant. Data was excluded from the correlation analyses if all three assessments were not completed by both coders.

### Interrater reliability

Interrater reliability was evaluated between the two primary assessors, a physical therapist and an occupational therapist, both with greater than 20 years of experience and both clinical experts in evaluating children with *STXBP1*-RD and *SYNGAP1*-RD. The order of assessments and time between assessments were based on the participant’s clinic schedule, which was determined by provider availability and therefore was unable to be standardized. If a participant was unable to be scored due to fatigue or behavior by both assessors for any individual classification scale, the data for that classification scale was not included in the analysis.

### Test-retest stability

Test-retest stability was evaluated for participants with at least two completed longitudinal visits. The functional rating scale scores were compared between the participant’s first and second assessment visits. If both raters completed the scales, the consensus score was used; if only one rater completed the scale, the score from that rater was used.

### Statistical analysis

Statistical analysis was completed using the R Statistical Framework. Data for the miniMACS and MACS were combined due to the limited number of subjects who were under four years of age. Percentage agreement and Cohen’s kappa statistic with squared weights were calculated as per CONSORT recommendations for assessing reliability.^26^ This method accounts for the degree of disagreement by penalizing larger differences more heavily than smaller ones. Inter-rater reliability was evaluated between the two primary coders while test-rest reliability was analyzed between visit 1 and 2. Weighted kappas were interpreted as follows: 0–.20 = no agreement, .21–.39 = minimal agreement, .40–.59 = weak agreement, .60–.79 = moderate agreement, and .80-.90 = strong agreement, above = .90 = almost perfect agreement.^27^ Kendall’s Tau was selected to evaluate the relations between the functional classification scales as it provides a robust measure of ordinal agreement that is less sensitive to ties and more appropriate for small sample sizes than Spearman’s rho.^28^ Statistical significance was set at p < .05. Kendall’s Tau was interpreted as follows: <0.10 = very weak association, 0.10–0.30 = weak association, 0.30–0.50 = moderate association, 0.50–0.70 = strong association, and >0.70 = very strong association.^29^

## Results

The sample of individuals with *STXBP1*-RD with GMFCS, MACS/mini-MACS, and CFCS scores included a total 83 individuals and consisted of 34 females and 49 males with a mean age of 9.8 years at the time of assessment with a range of 2.5-43.4 years. The sample of 101 individuals with *SYNGAP1*-RD consisted of 51 females and 50 males who had a mean age of 10.9 years with a range of 2.4-69.0 years. The mean time between assessment 1 and assessment 2 for the stability analysis was 9 months for both groups (range 4-16 months).

The classification scale scores for individuals with *STXBP1*-RD varied among domains (Table 1). A slight majority of GMFCS scores were either Levels I or II, most participants scored at Level II, III, or IV in MACS/miniMACS, and most CFCS scores were either Levels III or IV. The classification scale scores for individuals with *SYNGAP1-*RD were less variable among domains (Table 1) with over 95% and 69% of scores at either GMFCS or MACS/mini–MACS Levels I or II, respectively while 86% of participants scored at Level II, III, or IV in the CFCS (Figure 1). Compared to data reported by Palisano^25^ in children with CP (Table 1), individuals with *STXBP1*-RD had a lower percentage of children rated as a Level I on all three functional scales while individuals with *SYNGAP1*-RD had better gross motor function as measured by GMFCS ratings and worse language skills as measured by CFCS Level I ratings.

**Figure 1.**
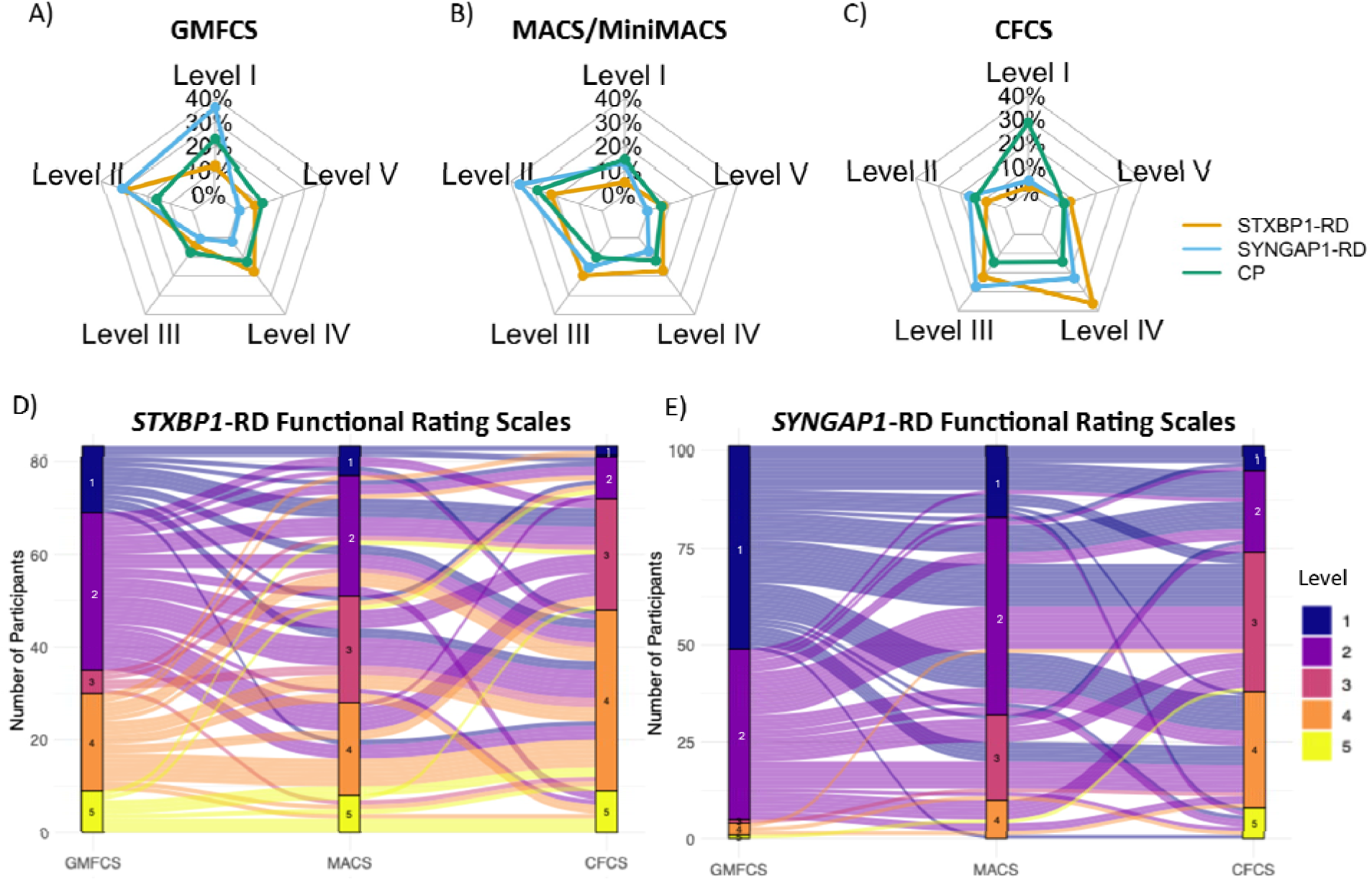
Distribution of scores by diagnosis and across functional scales. A-C) Distribution of severity levels among participants with *STXBP1*-RD, *SYNGAP1*-RD and a published cohort of individuals with CP.^25^ D-E) Distribution of participants with *STXBP1*-RD and *SYNGAP1*-RD between the GMFCS, MACS/MiniMACS, and CFCS functional rating scales. Abbreviations: *STXBP1*-Related Disorders (*STXBP1*-RD); *SYNGAP1*-Related Disorders (*SYNGAP1*-RD), Gross Motor Function Classification System (GMFCS); mini–Manual Ability Classification System (mini-MACS); Manual Ability Classification System (MACS); Communication Function Classification System (CFCS).

**Table 1:** Number of individuals with a classification score with percentage in parentheses for the first assessment visit for participants with *STXBP1*-RD, *SYNGAP*1-RD, and a published cohort of individuals with CP.^25^.

| ***STXBPI*-RD (n=83)** |  |  |  |
| --- | --- | --- | --- |
|  | GMFCS | MACS/MiniMACS | CFCS |
| Level I | 14 (16.9%) | 6 (7.2%) | 2 (2.4%) |
| Level II | 34 (41.0%) | 26 (31.3%) | 9 (11.1%) |
| Level III | 5 (6.0%) | 23 (27.7%) | 24 (28.9%) |
| Level IV | 21 (25.3%) | 20 (24.1%) | 39 (47.0%) |
| Level V | 9 (10.8%) | 8 (9.6%) | 9 (11.1%) |
| ***SYNGAP1*-RD (n=101)** |  |  |  |
|  | GMFCS | MACS/MiniMACS | CFCS |
| Level I | 52 (51.5%) | 18 (17.8%) | 6 (5.9%) |
| Level II | 44 (43.6%) | 51 (50.5%) | 21 (20.8%) |
| Level III | 1 (1.0%) | 22 (21.8%) | 36 (35.6%) |
| Level IV | 3 (3.0%) | 10 (9.9%) | 30 (29.7%) |
| Level V | 1 (1.0%) | 0 (0%) | 8 (7.9%) |
| **Cerebral Palsy (Palisano, 2018<sup>25</sup>) (n=652)** |  |  |  |
|  | GMFCS | MACS | CFCS |
| Level I | 213 (32.7%) | 131 (20.1%) | 242 (37.1%) |
| Level II | 148 (22.7%) | 259 (39.7%) | 115 (17.6%) |
| Level III | 72 (11.0%) | 94 (14.4%) | 124 (19.0%) |
| Level IV | 118 (18.1%) | 111 (17.0%) | 122 (18.7%) |
| Level V | 101 (15.5%) | 57 (8.7%) | 49 (7.5%) |

Abbreviations: *STXBPI*-Related Disorders (*STXBPI*-RD); *SYNGAP1*-Related Disorders (*SYNGAP1*-RD), Gross Motor Function Classification System (GMFCS); mini-Manual Ability Classification System (mini-MACS); Manual Ability Classification System (MACS); Communication Function Classification System (CFCS).

Interrater reliability and stability are presented in Table 2. Interrater agreement varied from 73.8% to 77.3% for the *STXBP1*-RD cohort and 60.5% to 83.3% for the *SYNGAP1*-RD cohort. Interrater reliability weighted kappas for the *STXBP1*-RD cohort were 0.83 to 0.93 while the *SYNGAP1*-RD cohort were 0.66-0.81. Test-retest stability scores for the *STXBP1*-RD group varied from 0.62 to 0.94 while the *SYNGAP1*-RD group varied from 0.38 to 0.78 (Figure 2) and the absolute difference between the raters is presented in Table 3. Correlations are presented in Table 2 while cross tabulations are presented in Table 4. The relations were statistically significant between all scales for both cohorts and Kendall’s Tau values ranged from 0.25 to 0.42 for the *STXBP1*-RD cohort and 0.19 to 0.45 for the *SYNGAP1*-RD cohort.

**Figure 2.**
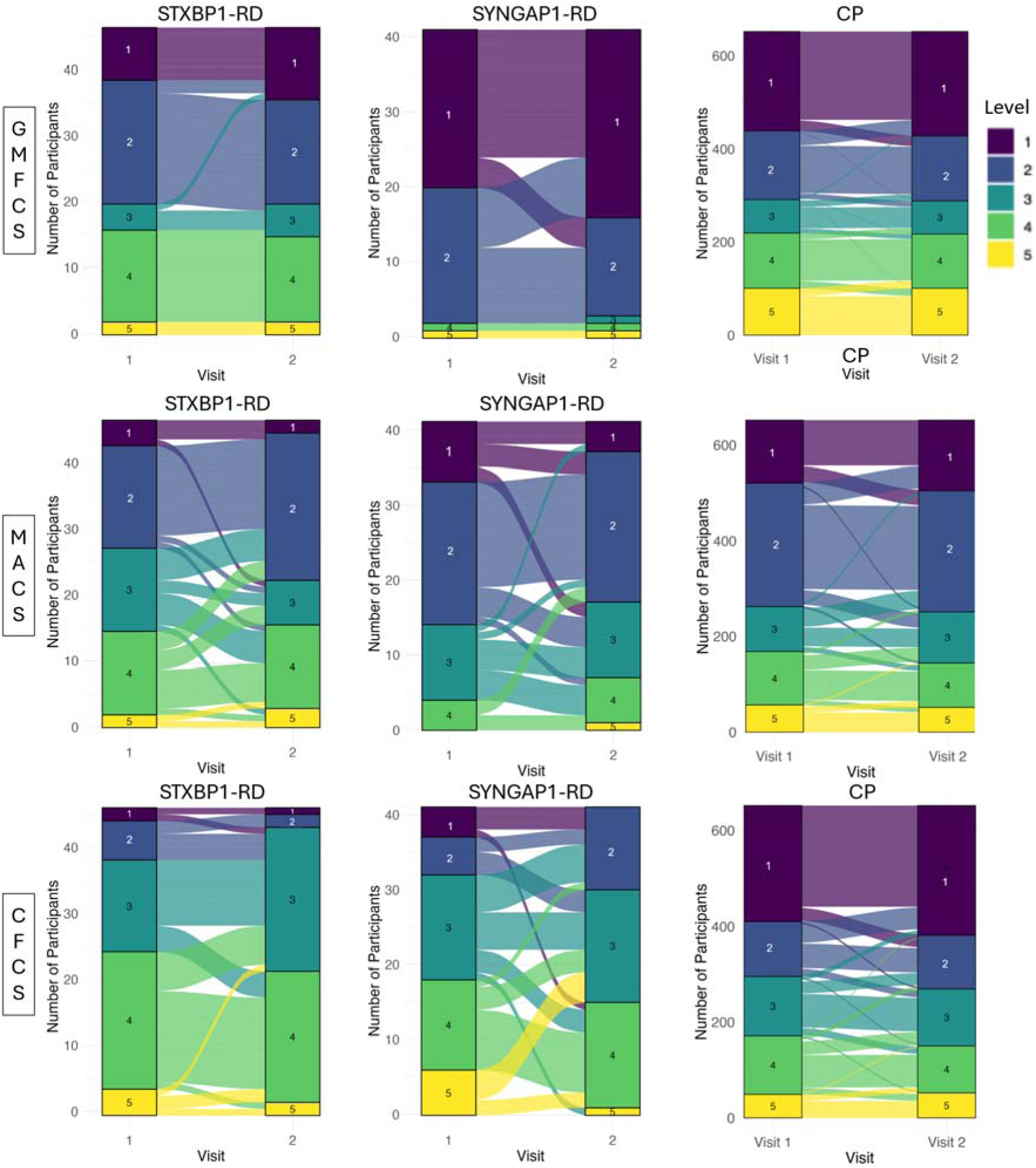
Test-Retest stability across time by diagnosis and functional rating scale. Abbreviations: *STXBP1*-Related Disorders (*STXBP1*-RD); *SYNGAP1*-Related Disorders (*SYNGAP1*-RD), Gross Motor Function Classification System (GMFCS); mini–Manual Ability Classification System (mini-MACS); Manual Ability Classification System (MACS); Communication Function Classification System (CFCS).

**Table 2:** Inter-rater Reliability, Test-Retest Stability, and Scale Correlations.

| **Inter-rater Reliability** |  |  |  |  |
| --- | --- | --- | --- | --- |
| **Diagnosis** | **Scale** | **n** | **Weighted Kappa** | **Percent Agreement** |
| *STXBPI*-RD | GMFCS | 44 | 0.928 | 77.3 |
|  | MACS/MiniMACS | 42 | 0.832 | 73.8 |
|  | CFCS | 42 | 0.831 | 76.2 |
| *SYNGAPI*-RD | GMFCS | 42 | 0.788 | 83.3 |
|  | MACS/MiniMACS | 41 | 0.655 | 63.4 |
|  | CFCS | 43 | 0.807 | 60.5 |
| **Test-Retest Stability** |  |  |  |  |
| **Diagnosis** | **Scale** | **n** | **Weighted Kappa** | **Percent Agreement** |
| *STXBPI*-RD | GMFCS | 47 | 0.942 | 89.4 |
|  | MACS/MiniMACS | 48 | 0.616 | 52.1 |
|  | CFCS | 47 | 0.637 | 59.6 |
| *SYNGAPI*-RD | GMFCS | 41 | 0.777 | 68.3 |
|  | MACS/MiniMACS | 41 | 0.477 | 53.7 |
|  | CFCS | 41 | 0.376 | 36.6 |
| **Correlations** |  |  |  |  |
| **Diagnosis** | **Scale Pairs** | **n** | **Kendall's Tau** | **p-value** |
| *STXBPI*-RD | GMFCS vs MACS/MiniMACS | 83 | 0.416 | <.001 |
|  | GMFCS vs CFCS | 83 | 0.251 | .007 |
|  | MACS/MiniMACS vs CFCS | 83 | 0.27 | .004 |
| *SYNGAPI*-RD | GMFCS vs MACS/MiniMACS | 101 | 0.453 | <.001 |
|  | GMFCS vs CFCS | 101 | 0.189 | .035 |
|  | MACS/MiniMACS vs CFCS | 101 | 0.335 | <.001 |

Abbreviations: *STXBPI*-Related Disorders (*STXBPI*-RD); *SYNGAPI*-Related Disorders (*SYNGAPI*-RD), Gross Motor Function Classification System (GMFCS); mini-Manual Ability Classification System (mini-MACS); Manual Ability Classification System (MACS); Communication Function Classification System (CFCS).

**Table 3:** Differences between Visit 1 and Visit 2 scores.

| **Diagnosis** | **Functional Rating Scale** | **Absolute Difference** | **Count** | **Percentage** |
| --- | --- | --- | --- | --- |
| *STXBPI*-RD | GMFCS | 0 | 42 | 89.4 |
|  |  | 1 | 4 | 8.5 |
|  |  | 2 | 1 | 2.1 |
| *STXBPI*-RD | MACS | 0 | 25 | 52.1 |
|  |  | 1 | 17 | 35.4 |
|  |  | 2 | 6 | 12.5 |
| *STXBPI*-RD | CFCS | 0 | 28 | 59.6 |
|  |  | 1 | 17 | 36.2 |
|  |  | 2 | 2 | 4.3 |
| *SYNGAPI*-RD | GMFCS | 0 | 28 | 68.3 |
|  |  | 1 | 13 | 31.7 |
| *SYNGAPI*-RD | MACS | 0 | 22 | 53.7 |
|  |  | 1 | 13 | 31.7 |
|  |  | 2 | 6 | 14.6 |
| *SYNGAPI*-RD | CFCS | 0 | 15 | 36.6 |
|  |  | 1 | 19 | 46.3 |
|  |  | 2 | 6 | 14.6 |
|  |  | 3 | 1 | 2.4 |

Abbreviations: *STXBPI*-Related Disorders (*STXBPI*-RD); *SYNGAPI*-Related Disorders (*SYNGAPI*-RD), Gross Motor Function Classification System (GMFCS); mini-Manual Ability Classification System (mini-MACS); Manual Ability Classification System (MACS); Communication Function Classification System (CFCS).

**Table 4.** Cross Tabulations.

| **Gross Motor Function Classification System** |  |  |  |  |  |  |
| --- | --- | --- | --- | --- | --- | --- |
| *STXBPI-*<br>*RD* | **Rater 2** |  |  |  |  |  |
| **Rater 1** | **I** | **II** | **III** | **IV** | **V** | **Total** |
| **I** | 6 | 1 | 0 | 0 | 0 | 7 |
| **II** | 2 | 15 | 2 | 0 | 0 | 19 |
| **III** | 0 | 0 | 5 | 0 | 0 | 5 |
| **IV** | 0 | 0 | 3 | 10 | 1 | 14 |
| **V** | 0 | 0 | 0 | 1 | 3 | 4 |
| **Total** | 8 | 16 | 10 | 11 | 4 | 49 |
| *SYNGAP*<br>*I-RD* | **Rater 2** |  |  |  |  |  |
| **Rater 1** | **I** | **II** | **III** | **IV** | **V** | **Total** |
| **I** | 13 | 2 | 0 | 0 | 0 | 15 |
| **II** | 4 | 21 | 0 | 0 | 0 | 25 |
| **III** | 0 | 0 | 0 | 0 | 0 | 0 |
| **IV** | 0 | 1 | 0 | 0 | 0 | 1 |
| **V** | 0 | 0 | 0 | 0 | 1 | 1 |
| **Total** | 17 | 24 | 0 | 0 | 1 | 42 |

| **Manual Ability Classification System and Mini MACS** |  |  |  |  |  |  |
| --- | --- | --- | --- | --- | --- | --- |
| *STXBPI-*<br>*RD* | **Rater 2** |  |  |  |  |  |
| **Rater 1** | **I** | **II** | **III** | **IV** | **V** | **Total** |
| **I** | 3 | 0 | 1 | 0 | 0 | 4 |
| **II** | 0 | 10 | 0 | 0 | 0 | 10 |
| **III** | 0 | 3 | 12 | 4 | 1 | 20 |
| **IV** | 0 | 1 | 0 | 6 | 2 | 9 |
| **V** | 0 | 0 | 0 | 1 | 3 | 4 |
| **Total** | 3 | 14 | 13 | 11 | 6 | 47 |
| *SYNGAP*<br>*I-RD* | **Rater 2** |  |  |  |  |  |
| **Rater 1** | **I** | **II** | **III** | **IV** | **V** | **Total** |
| **I** | 5 | 2 | 0 | 0 | 0 | 7 |
| **II** | 3 | 10 | 0 | 0 | 0 | 13 |
| **III** | 0 | 2 | 7 | 1 | 0 | 10 |
| **IV** | 1 | 2 | 2 | 5 | 0 | 10 |
| **V** | 0 | 0 | 0 | 1 | 0 | 1 |
| **Total** | 9 | 16 | 9 | 7 | 0 | 41 |

| Communication Function Classification System |  |  |  |  |  |  |
| --- | --- | --- | --- | --- | --- | --- |
| ***STXBPI-***<br>***RD*** | **Rater 2** |  |  |  |  |  |
| **Rater 1** | **I** | **II** | **III** | **IV** | **V** | **Total** |
| **I** | 1 | 0 | 0 | 0 | 0 | 1 |
| **II** | 0 | 3 | 0 | 0 | 0 | 3 |
| **III** | 0 | 1 | 9 | 2 | 0 | 12 |
| **IV** | 0 | 1 | 1 | 14 | 3 | 19 |
| **V** | 0 | 0 | 0 | 2 | 5 | 7 |
| **Total** | 1 | 5 | 10 | 18 | 8 | 42 |
| ***SYNGAP***<br>***I-RD*** | **Rater 2** |  |  |  |  |  |
| **Rater 1** | **I** | **II** | **III** | **IV** | **V** | **Total** |
| **I** | 3 | 1 | 0 | 0 | 0 | 4 |
| **II** | 1 | 4 | 2 | 0 | 0 | 7 |
| **III** | 0 | 3 | 9 | 3 | 0 | 15 |
| **IV** | 0 | 1 | 3 | 6 | 0 | 10 |
| **V** | 0 | 0 | 0 | 2 | 4 | 6 |
| **Total** | 4 | 9 | 14 | 11 | 4 | 42 |

## Discussion

Our investigation reported GMFCS, MACS/MiniMACS, and CFCS scores in the largest sample of individuals with *STXBP1*-RD to date and is the first to use these scales in individuals with *SYNGAP1*-RD. The use of classification scales designed for children with CP in populations with rare diseases such as *STXBP1*-RD and *SYNGAP1*-RD is likely to expand as the rate of diagnosis increases with the expansion of genetic testing. In addition, the development of gene and drug therapies for these conditions is ongoing, and future clinical trials may utilize these scales to describe and stratify participant phenotypes.^14,15^

In individuals with *STXBP1*-RD, our investigation found that language function was more impacted than gross motor function. These results support previous research that described a similar pattern of functional performance.^21,22^ Additionally, families also have reported more concerns about language than gross motor skills.^9^ However, our sample had improved gross motor function compared to Thalwitzer et al.^22^ Most of our individuals with *STXBP1*-RD had GMFCS levels I and II while 59% of individuals reported by Thalwitzer et al^22^ had GMFCS Levels IV and V. These results may be due to the larger sample size of our natural history study, differences in patient populations between countries due to referral differences for genetic testing, and due to retrospective assignment of GMFCS levels by Thalwitzer. While Xian et al^21^ reported MACS scores in a small subset of nine individuals, our study is the first to systematically report MACS/miniMACS scores in a large sample of subjects. A similar pattern of greater limitations in language compared to gross and fine motor function was observed in individuals with *SYNGAP1*-RD. To date, there is limited literature published on gross and fine motor function in *SYNGAP1*-RD since the phenotype is most associated with autism spectrum disorder, intellectual disability, and behavioral challenges.^13^ However, delays in the acquisition of gross motor, fine motor, and language milestones have been reported^13^ while Bednarczuk et al found that children with *SYNGAP1*-RD were more likely to have fine motor and language deficits compared to children with other causes of developmental delay.^24^ Future investigations with larger sample sizes using the GMFCS, MACS/MiniMACS, and CFCS are required to better define the functional abilities of individuals with *STXBP1*-RD and *SYNGAP1*-RD.

Our investigation is the first to assess the reliability and stability of the GMFCS, MACS/MiniMACS, or CFCS in any rare neurodevelopmental disorder. For individuals with *STXBP1*-RD, strong to almost perfect agreement with interrater reliability was found with kappa coefficients above .80. The kappa coefficients of individuals with *SYNGAP1*-RD were lower but still considered moderate to strong agreement, which may be due to the behavioral phenotype of *SYNGAP1*-RD leading to more variable function during assessments. Our results are similar to reliability studies using these scales with children with CP which found that these scales demonstrate moderate to strong reliability.^24^ Therefore, we suggest that these scales demonstrate appropriate levels of interrater reliability for use in clinical practice and research environments with individuals with *STXBP1*-RD and *SYNGAP1-*RD.

In our *STXBP1*-RD cohort, the stability of these scales was moderate (MACS/miniMACS) to almost perfect (GMFCS), which provides evidence for their use to track function over time. However, in our *SYNGAP1*-RD cohort only the GMFCS demonstrated moderate stability while the MACS/miniMACS and CFCS showed minimal or weak stability respectively. For both conditions, stability of scores were decreased when compared to the CP literature which found all three measures were more stable.^25^ The cause of this decreased stability is unknown, but we would hypothesize that the developmental trajectories of children with *STXBP1*-RD and *SYNGAP1*-RD may differ from those for children with CP especially since their phenotypes are different as shown by our classification scale comparison in Table 1. A second hypothesis for the decreased stability is that individuals with *STXBP1*-RD and *SYNGAP1*-RD often have drug resistant epilepsy which leads to variability in seizure control and subsequent variability in functional impairments. Additional research examining the stability of these scales in *STXBP1*-RD and *SYNGAP1*-RD, which may be completed during natural history studies, is necessary to further explore these hypotheses.

While relations between all scales reached statistical significance for both cohorts, the strength of the correlations ranged from very weak to moderate. This finding differs from the CP literature in which strong positive correlations between the GMFCS, MACS/MiniMACS, or CFCS have been reported.^23^ Since both cohorts demonstrate higher levels of gross motor function compared to language function as measured by the functional rating scales, a weaker correlation may be expected. In addition, the overall lack of variability of scores in both samples may lead to lower correlations. Future investigations with larger sample sizes and more functionally diverse samples may be useful in providing additional information on the relations between these scales in populations with *STXBP1*-RD and *SYNGAP1*-RD.

Despite the large sample size, we needed to combine age-groups for the functional rating scales, including the MACS and miniMACS, and GMFCS for analysis. There were a limited number of participants under 4 years given that many individuals with *STXBP1*-RD and *SYNGAP1*-RD do not receive a molecular diagnosis prior to this age. In addition, due to the lack of standardized classification tools for adults, out of age range use of the scales was completed which commonly occurs in rare conditions.^30^ Additional research is needed to determine the validity and reliability of these classification scales in adults with *STXBP1*-RD and *SYNGAP1*-RD.

## Conclusion

We provide evidence that the GMFCS, MACS/MiniMACS, and CFCS can be reliably assessed in individuals with *STXBP1*-RD and *SYNGAP1*-RD and that language function is more impaired than gross and fine motor function in both conditions. Weak correlations were found between scales in both populations while the stability of these scales over time is less evident. The functional rating scales assessed in our study can be used to inform care, aid in identifying subgroups based on phenotypic features, and to classify individuals in future intervention trials.

## Data Availability

All data produced in the present study are available upon reasonable request to the authors

## Acknowledgements

This study was supported by The Center for Epilepsy and Neurodevelopmental Disorders (ENDD), the National Institute for Neurological Disorders and Stroke (R01 NS127830-01A1 and R01 NS131512-01 IH), the American Epilepsy Society (AES), Pediatric Epilepsy Research Foundations (PERF) & SynGAP Research Fund (SRF) through a Research Training Fellowship for Clinicians (JLM), and the American Academy of Neurology (AAN), AES, the Epilepsy Foundation, & the American Brain Foundation (ABF) through the Susan Spencer Award (JLM). SMR is supported by an individual grant from the STXBP1-disorders foundation and by the Career Ladder Education Program for Genetic Counselors grant from the Warren Alpert Foundation.

## Notes

### Competing Interest Statement

The authors have declared no competing interest.

### Author Declarations

IRB of Children’s Hospital of Philadelphia gave ethical approval for this work

